# Hybrid immunity versus vaccine-induced immunity against SARS CoV2 in Patients with Autoimmune Rheumatic Diseases

**DOI:** 10.1101/2021.08.26.21258418

**Authors:** Padmanabha Shenoy, Sakir Ahmed, Aby Paul, Somy Cherian, Rashwith Umesh, Veena shenoy, Anuroopa Vijayan, Sageer Babu, S Nivin, Arya Thambi

## Abstract

**Introduction:** Single-dose COVID-19 vaccines in healthy individuals with past COVID-19 infections seem to provide better immunity than double doses in COVID-19 unexposed individuals. However, it is not known whether the same is true for patients with autoimmune rheumatic diseases (AIRD) who are on immunosuppressants.

**Methods:** We identified 30 patients with AIRD who took a single dose of the ChAdOx1 vaccine post-COVID-19 infection. Age, sex and disease similar patients were enrolled in to three groups of 30 each who had (1) past infection with COVID-19 but no vaccine, (2) a single dose of ChAdOx1 and (3) double doses of ChAdOx1. Sera were collected from each patient approximately 30 days after last vaccine dose or since the onset of COVID19 symptoms (in the unvaccinated group). Antibodies to spike protein were estimated and virus neutralization potential of sera was tested.

**Results:** Baseline characteristics including drug usage was similar betweenthe groups. Seroconversion occurred in 25(83%), 23(77%), 27(90%), and 30(100%) in natural infection, single-dose vaccine, double dose vaccine, and infection +single dose vaccine groups respectively. Mean antibody titres (10076.8±8998) in the last group were at least 6-100x higher than in the other 3 groups. Also, the infection +vaccine group had the highest neutralization potential of 83.37 % as compared to 45.4% in the fully vaccinated group.

**Conclusion:** The hybrid immunity with a single dose of the vector-based vaccine post-infection seems to be superior to double dosage of the vaccine in patients with AIRD. A universal vaccination strategy involving a single dose of vaccine for all individuals with previous COVID-19 infection seems to be effective in these patients also.

**What is already known about this subject?:** A single dose of an RNA based COVID-19 vaccine after COVID-19 natural infection provides superior immune protection as compared to double doses of vaccines in infection naïve persons

A second dose of vaccine in healthy people who had infection previously does not increase the immune protection but may paradoxically induce tolerance

Vaccine responses in patients with autoimmune rheumatic diseases(AIRD) may be suboptimal due to underlying disease or the use of immunosuppressants.

**What does this study add?:** Hybrid-induced immunity (single vaccine post COVID-19 infection) produces adequate vaccine responses in patients with AIRD, non-inferior to double dose of vaccine

Besides mRNA vaccines, the adenoviral vector vaccine AZD1222 also demonstrates this hybrid phenomenon.

**How might this impact on clinical practice?:** Vaccination policies can consider providing only a single vaccine in those who had previous COVID-19 infection. This strategy has been shown not to be harmful for patients with AIRD. This will help reduce vaccine shortages.

## Introduction

Vaccination is currently the strongest weapon in our fight against the COVID-19 pandemic. The World Health Organisation is warning about the third major wave across the globe[1]. As long as the entire globe is not covered with adequate vaccination, there is always a danger of SARS-CoV-2 strains spreading fast in susceptible populations leading to rapid mutations and selection of more virulent and vaccine-escape mutants[2,3]. Thus, there needs to be adequate and equitable vaccine distribution throughout the world. Understanding vaccine safety and immunogenicity are of paramount importance to facilitate vaccine policies.

Patients with autoimmune rheumatic diseases (AIRD) are a special group in whom the virus may stay for prolonged periods and this too has been hypothesized to lead to newer virus variants[4,5]. With the reports of weaning of immunity after 6 months from many parts of the world, a third dose is being administered by a few countries like Israel to their high-risk population[6]. The United States Food and Drug Administration has approved a third dose for immunocompromised patients [7]. This third dose will add to the vaccine supply chain constraints that the world is already facing. Public health policies backed by scientific evidence is urgently needed to mitigate the dearth of vaccines.

There have been reports that a single dose of mRNA vaccine in people with a history of COVID infection induces an immune response similar to those who have received two doses of vaccine and no history of COVID infection [8–11]. Similar data have been put forward for the vector-borne vaccine ChAdOx1 in a single study[12]. Overall limited data isavailable comparing the immunogenicity of vector-based vaccines in those with prior infection and those who have received two doses without infection. We are not aware of such reports in patients with AIRD.

Patients with AIRD has been shown to have a suboptimal response to vaccines[13,14]. Most immunosuppressive drugs appear to have moderate to no effect on immunogenicity. However, drugs like Rituximab have also been shown to be associated with lower rate of seroconversion post-vaccination[15]. We have previously shown that patients on immunosuppressive therapy respond similarly to the healthy controls following SARS-CoV-2 infection[16]. A small proportion of patients may experience flares of their autoimmune diseases following vaccination. It is important to understand the immune response of a single dose of vaccine in immunosuppressed persons with a history of COVID-19 infection and compare it with those who have received two doses without any history of COVID-19.

We are following up a cohort of around 1500 patients with AIRD who have received COVID-19 vaccines. Out of this cohort, we sought to compare the patients who had natural infections only or natural infection with one dose of the ChAdOx1 vaccine with those who have completed either one or both doses of the ChAdOx1 vaccine. We aimed to look at the safety and immunogenicity of vaccines post-natural COVID-19 infections, including the neutralization potential of antibodies produced by natural infections.

## Methods

Our cohort comprises patients with AIRD registered with our centre (CARE – Centre for Arthritis and Rheumatism Excellence). We have been following up AIRD patients who have had COVID-19 infections [16] and those who have received vaccines [17]. Serum and plasma samples of these patients have been stored for future research with patients’ informed consent. We identified 30 patients who had documented COVID-19 infectionin the last 12 months and have received one dose of the ChAdOX1 vaccine (Group IV). We took age and sex-matched patients (30 in each group) receiving similarimmunosuppressants havingeither: (1) documented COVID-19 in the last 6 months but has still not received any vaccine(Group I); (2) received a single dose of the ChAdOx1 vaccine only (Group V) and (3) completed two doses of ChAdOx1 vaccination as per prevalent national guidelines(Group VV). In the last two groups, patients were queried for any symptoms suggestive of COVID-19 in the past and they were excluded from vaccine only groups if they reported any such symptoms. The most commonly used vaccine in India is the ChAdOx1 ncov-19 vaccine (AZD1222), a non replicating chimpanzee adenovirus vector carrying the SARS-COV2 structural surface glycoprotein antigen (spike protein) gene. To avoid confounding due to different vaccines, we have included patients who were administered the ChAdOx1 vaccine only.

Serum was collected between 4-6 weeks of receiving the last vaccine dose from the patients who had received any vaccine. For patients with only natural infection, serum was collected approximately 30 days after the date of first symptoms. These were being routinely done for all our AIRD patients post-infection and post-vaccination after informed consent as a part of our larger COVID-19 vaccination in the AIRD study. All sera and plasma were stored in aliquots at -80° C until processing.

Quantitative measurement of IgG Antibodies to S1 protein of the SARS-CoV-2 virionwas done using the Elecsys Anti-SARS-CoV-2 Chemiluminescent assay (Roche, Switzerland). Viral neutralization assay was assessed by SARS-CoV-2 sVNT Kit (GenScript). This kit detects Neutralising Ab that blocks the interaction between the angiotensin-converting enzyme 2 cell surface receptor and RBD of the SARS CoV-2 Spike protein. If the inhibition ≥30%, sample was classified as “positive”.

The normality of data was checked by the Shapiro-Wilk test. The difference in proportions was tested by the Fisher Exact test and the difference in antibody titres was tested by Independent sample t-test after log transformation. P<0.05 was the cut-off for significance. For multiple comparisons, p-values were adjusted as per Benjamin Hochberg corrections. All analysis and graphical representations were carried out in R version 3.4 using various packages including ggplot2, ggforce, Hmisc, rstatix, and rocr.

Ethics approval for the study was obtained from Sree Sudheendra Medical mission (IEC/2021/35) Informed consent was obtained from each patient before sample collection.

## Results

Of the total of 120 patients in the 4 groups, average age was 55.82 and (88.33%) were females. Rheumatoid arthritis was the most common rheumatic disease. Baseline characteristics including drugs used were similar amongst the 4 groups with 30 patients each [Table 1].

**Table 1:** Comparison of baseline characteristics between the four groups (30 patients of autoimmune rheumatic disease each).

| Group |  | V | I | VV | VI | P value |
| --- | --- | --- | --- | --- | --- | --- |
| Description |  | Single dose Vaccine (n=30) | COVID Infection Only (n=30) | Double dose vaccine (n=30) | Infection + Single dose vaccine (n=30) | Significance |
| Age (Mean ±SD) | | 56.77±9.28 | 55.73±8.28 | 56.07±8.15 | 54.73±8.02 | 0.825 |
| Females |  | 28(93.33%) | 27(90%) | 26(86.66%) | 25(83.33%) | 0.655 |
| Diseases | Rheumatoid arthritis | 18(60%) | 18(60%) | 24(80%) | 21(70%) | 0.288 |
|  | Spondyloarthropathy | 4(13.33%) | 2(6.66) | 2(6.66) | 2(6.66) | 0.725 |
|  | Systemic lupus erythematosus | 2(6.66) | 5(16.66%) | 1(3.33%) | 2(6.66) | 0.283 |
|  | Others | 6(20%) | 5(16.66%) | 3(10%) | 5(16.66%) | 0.756 |
| Methotrexate |  | 18(60%) | 18(60%) | 15(50%) | 17(56.66%) | 0.887 |
| Hydroxychloroquine |  | 19(63.33%) | 19(63.33%) | 23(76.66%) | 23(76.66%) | 0.616 |
| Sulfasalazine |  | 4(13.33%) | 4(13.33%) | 4(13.33%) | 3(10%) | 0.973 |
| Leflunomide |  | 3(10%) | 3(10%) | 3(10%) | 3(10%) | 0.956 |
| Mycophenolate mofetil |  | 3(10%) | 3(10%) | 3(10%) | 3(10%) | 1 |
| Azathioprine |  | 0 | 0 | 0 | 0 | 0.795 |
| Steroid |  | 8(26.66%) | 8(26.66%) | 8(26.66%) | 6(20%) | 0.731 |

Group IV (Single dose vaccine after natural infection) had the highest antibody titre compared with all the three other groups (p>0.0001) [Figure 1]. Most of the antibody titres in Group IV (infection plus vaccine) is more than 1000, while it was less than 1000 in both the natural infection (I), 2 dose vaccines (VV) and single-dose vaccine (V) groups. The antibody titres, seroconversion rates and neutralization percentages in the four groups are summarized in table 2. There was 100% seroconversion in the IV group that was numerically more than in the fully vaccinated VV group (90%; p=>0.0001).

**Table 2:** Antibody titres and neutralization assays in the four groups (30 patients of autoimmune rheumatic disease each).

| Group | V | I | VV | VI |
| --- | --- | --- | --- | --- |
| Description | Single dose Vaccine (n=30) | COVID Infection Only (n=30) | Double dose vaccine (n=30) | Single dose vaccine + Infection (n=30) |
| Antibody titres Median (IQR) | 20.30(2.93-49.67) | 87.75(15.95-401.40) | 322.60(66.17-2353.75) | 11144.50(3392-20437.75) |
| Seroconversion (%) | 23 (76.7%) | 25 (83.3%) | 27 (90%) | 30 (100%) |
| %age neutralization of virion particles Median (IQR) | 5.91(0-16.01) | 47.75(3.36-84.89) | 35.71(2.34-89.48) | 83.37(57.81-92.35) |
| Proportion showing at least 30% neutralization | 5 (16.7%) | 16 (53.3%) | 18 (60%) | 26 (86.7%) |

**Figure 1:**
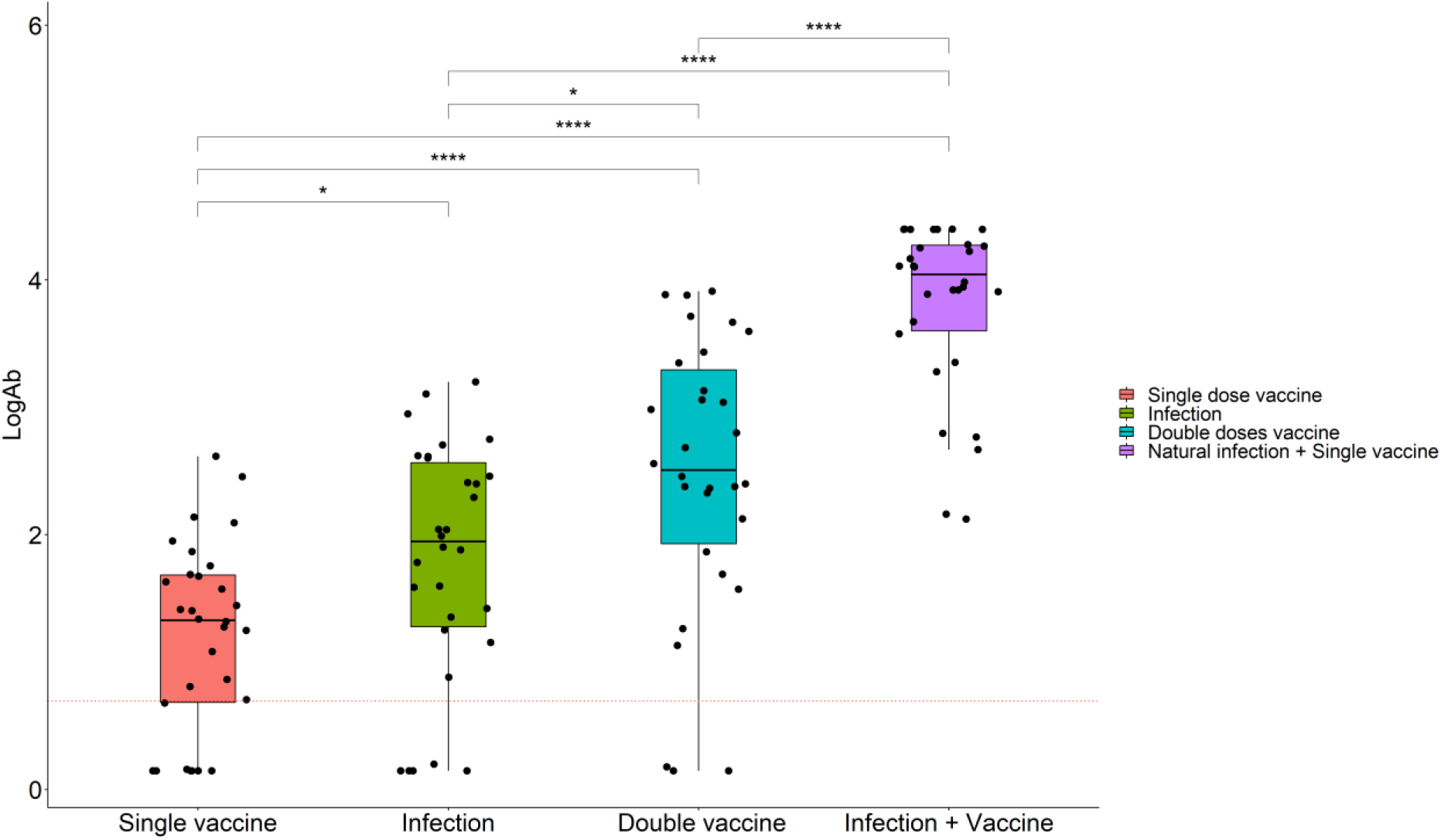
Box plot showing antibody levels in the 4 groups: Single dose Vaccine (n=30); COVID Infection Only (n=30); Double dose vaccine (n=30); and Single dose vaccine + Infection (n=30). P-values after correction for multiple comparisons: * p<0.05; *** p<0.001. The dotted red line represents the threshold for antibody positivity.

The proportion of patients in Group IV who had adequate neutralization(86.7%) was higher than that in the Group VV (60%; P=0.039). Also, a greater proportion in the VI group had more than 75% neutralization, mirror the high antibody titres. The neutralization assay showed a moderate correlation (Pearson R= 0.35; P<0.001) with antibody titres (Figure 2). Adequate neutralization (>30% neutralization) was significantly associated with higher antibody titres (p<0.0001).A ROC analysis revealed that antibody titres above 212 predicted more than 30% neutralization with a sensitivity of 81.5% and a specificity of 83.6% [Supplementary figure 1].

**Figure 2:**
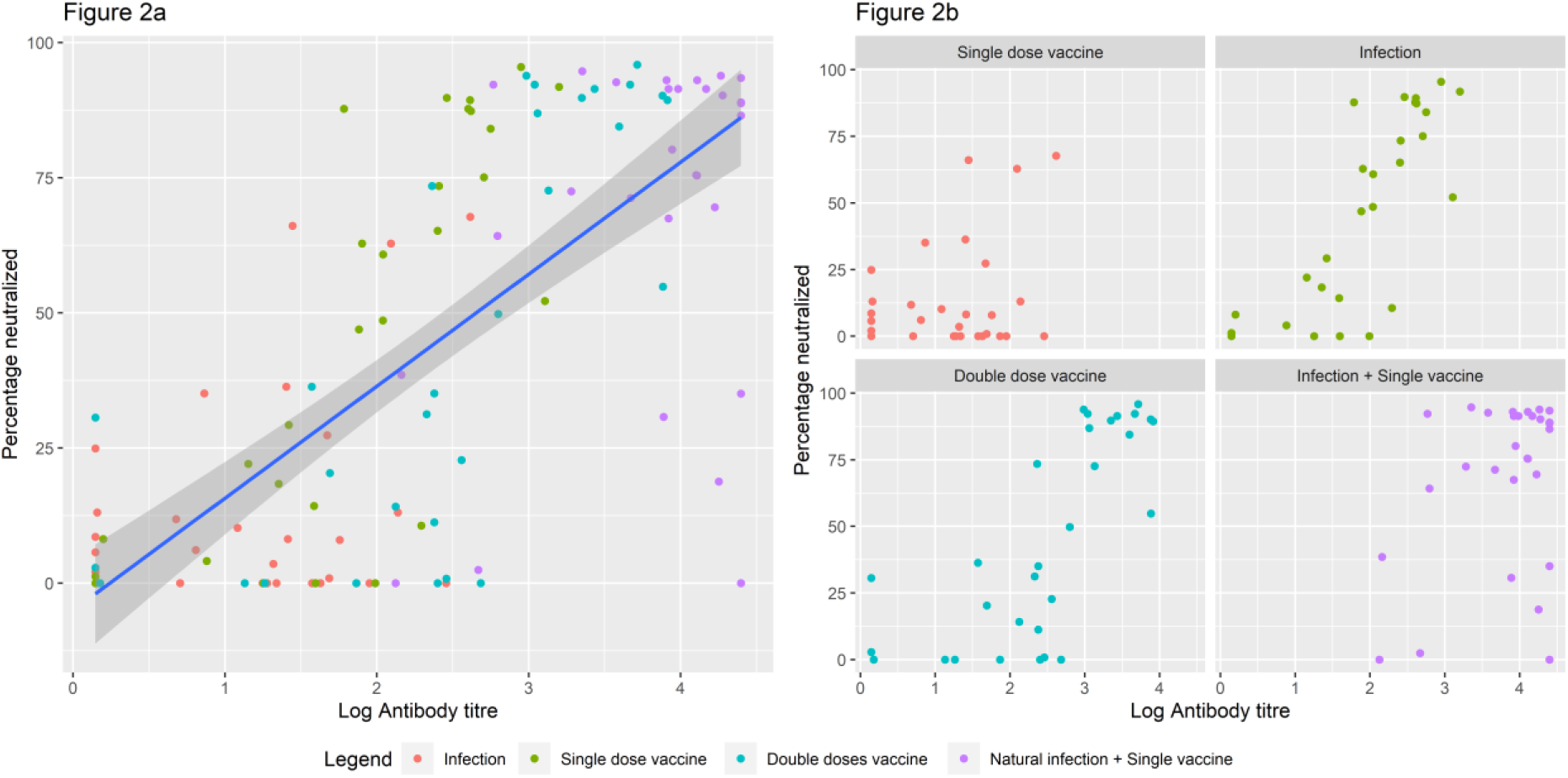
Scatter plot showing the relationship between absolute anti-Spike protein antibody levels and neutralization potential of the sera. (a) Overall in 120 patients and (b) divided as per groups. The navy blue line represents the regression line along with ± 2SD (dark grey area). Note the high neutralization potential as well as high antibody titres in the Infection + Vaccine group (purple colour).

## Discussion

We compared the serum antibody titres and viral neutralization assay in four groups of AIRD patients: documented COVID-19 infection, single-dose vaccinated, double dose vaccinated or single-dose vaccinated post-natural infection. Antibody titters were the highest in the infection with single vaccine (VI) group, at least ten to fifty-fold higher than that of any other group. There was 100% seroconversion and neutralization was also the highest in this group with 86.7% of sera showing more than 30% neutralization potential. This shows that a single dose of vaccine after COVID-19 natural infection provides better humoral immunity than two doses of a vaccine even in patients with AIRD.

We had previously shown that patients with AIRD also form adequate humoral responses to COVID-19 infection equivalent to those in healthy controls[16]. The current data shows that this can be augmented with a single dose of vaccine to produce a strong humoral immune response. A major concern was whether a natural infection would produce specific neutralizing antibodies. Hence, we tested in vitro the neutralizing capacity of the sera of these patients and found that antibodies produced are those with neutralising capability.

There have been multiple reports in small cohorts about the effectiveness of a single dose of an RNA based vaccine in patients who had COVID-19 before[8,10–12,18]. Most of these studies had been carried out in health care workers. Results similar to those of RNA vaccines have been found for the ChAdOx1 or AZD1222 vaccine in healthy individuals [9]. Though individually small, the combined interpretation of all these studies can be strong evidence in favour of such a strategy and it is being strongly recommended [19]. This concept of “hybrid immunity” induction is being explored in the context of vaccination post-natural infection [20]. The main concern in such a strategy would be whether it is sufficient for immunocompromised patients such as patients with AIRD.

We could show a robust hybrid immune response even in patients with AIRD. This has important implications for vaccination policies. With a worldwide shortage of vaccines, especially in low and middle-income countries, this provides an alternative to conserve vaccines. Such a policy of single vaccine post-natural infection is not likely to harm patients with autoimmune diseases or those on immunosuppressants. Moreover, there is a possibility that each vaccine dose predisposes the individual to the risk of an adverse event. Patients with AIRD are susceptible to flares due to repeated exposure to adjuvants present in vaccines. If they already have a strong immune response post-infection and a single dose of vaccine, an additional dose may predispose patients to more adverse effects including a flare of their underlying disease.

Individuals with prior exposure to SARS-CoV-2 demonstrated strong humoral and antigen-specific responses to the first dose but muted responses to the second dose of the vaccine[11]. A second dose may also prove to induce some amount of tolerance after such a high immune response.[21] This theoretically occurs due to T cell exhaustion post hyperstimulation and has been demonstrated in various viral infections[22].

The major strength of this study is that we had collected sera and plasma at approximate the same time (30 days) post-vaccination and post-infection. Also, the background disease and immunosuppression use are similar between the groups. The major limitation is that we have not looked at T cell immunogenicity. However, it has already been shown that peak neutralizing antibody titres correlates very well with the protection from COVID-19 infection probably independent of T cell responses[23].

Thus, in patients with AIRD who have had COVID-19 before, having a single dose of vaccine provides 10-50 fold higher humoral immunity than two doses of vaccine in infection naïve patients. This concept of hybrid immunity needs to be recognised and utilized in planning vaccination policies.

## Data Availability

Data available on request

## Patient and public involvement

Patients were not involved in the design, conduct or dissemination of the study

## Ethics approval

Ethics approval for the study was obtained from Sree Sudheendra Medical mission (IEC/2021/35)

## Contributorship

PS and SA conceptualised and drafted the paper. All authors approved the final version.

## Legends

Supplementary figure 1:

