## Supplementary figures and images for "Hybrid immunity versus vaccine-induced immunity against SARS CoV2 in Patients with Autoimmune Rheumatic Diseases"

### Supplementary figure 1

Supplementary Figure 1

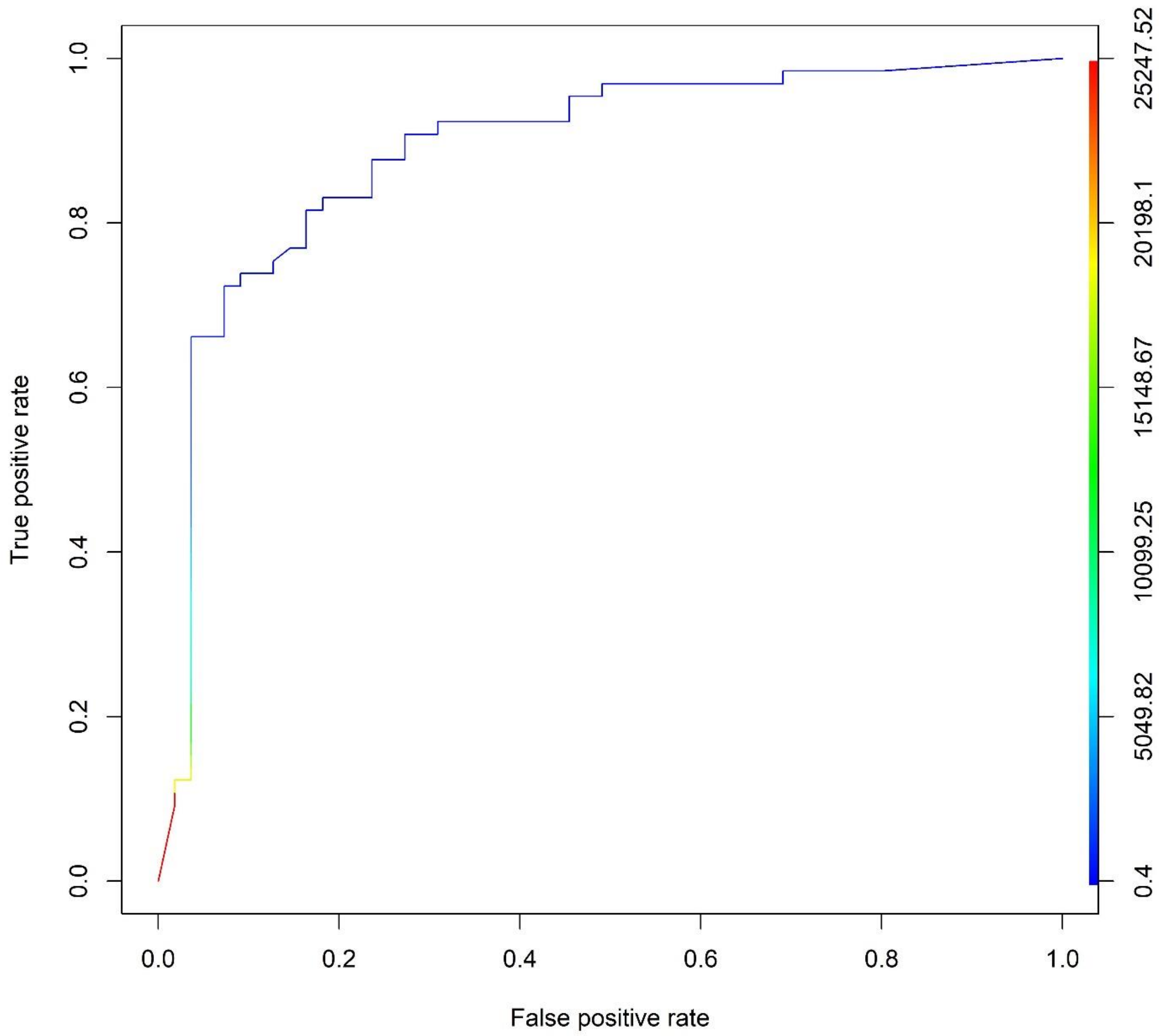
